# Optimizing Social Distancing Policies: A Dynamic Programming Approach for Coupled High and Low Risk Populations

**DOI:** 10.1101/2021.10.24.21265170

**Authors:** Peng Dai, Raffaele Vardavas, Sarah A. Nowak, Sze-chuan Suen

## Abstract

**Background:** Decision makers may use social distancing to reduce transmission between risk groups in a pandemic scenario like Covid-19. However, it may result in both financial, mental, and social costs. Given these tradeoffs, it is unclear when and who needs to social distance over the course of a pandemic when policies are allowed to change dynamically over time and vary across different risk groups (e.g., older versus younger individuals face different Covid-19 risks). In this study, we examine the optimal time to implement social distancing to optimize social utility, using Covid-19 as an example.

**Methodology:** We propose using a Markov decision process (MDP) model that incorporates transmission dynamics of an age-stratified SEIR compartmental model to identify the optimal social distancing policy for each risk group over time. We parameterize the model using population-based tracking data on Covid-19 within the US. We compare results of two cases: allowing the social distancing policy to vary only over time, or over both time and population (by risk group). To examine the robustness of our results, we perform sensitivity analysis on patient costs, transmission rates, clearance rates, mortality rates.

**Results:** Our model framework can be used to effectively evaluate dynamic policies while disease transmission and progression occurs. When the policy cannot vary by subpopulation, the optimal policy is to implement social distancing for a limited duration at the beginning of the epidemic; when the policy can vary by subpopulation, our results suggest that some subgroups (older adults) may never need to socially distance. This result may occur because older adults occupy a relatively small proportion of the total population and have less contact with others even without social distancing.

**Conclusion:** Our results show that the additional flexibility of allowing social distancing policies to vary over time and across the population can generate substantial utility gain even when only two patient risk groups are considered. MDP frameworks may help generate helpful insights for policymakers. Our results suggest that social distancing for high-contact but low-risk individuals (e.g., such as younger adults) may be more beneficial in some settings than doing so for low-contact but high-risk individuals (e.g., older adults).

## Introduction

Disease control has long relied on behavior modification techniques to reduce infection spread. At early stages of a pandemic, when interventions such as vaccination, antivirals, and antibiotics are not available, health officials must rely on implementing non-pharmaceutical interventions. These measures include social distancing, encouraging sick individuals to rapidly seek care, promoting hand washing and mask-wearing, or in more extreme cases, self-quarantining, among others. While these efforts may reduce disease spread, they may come at a high financial and psychological cost. For instance, to mitigate the spread of Covid-19, many countries have turned to social distancing, shutting nearly a hundred thousand businesses [1], separating family members [2] [3], and limiting social contact. The US lost 3.5 trillion dollars in GDP over the past year [4], had record levels of unemployment [5], and many suffered from the psychological toll and social stress due to distancing from friends and family.

Social distancing measures have generally taken the form of blanket guidelines intended to limit the total number of infections, rather than specifically limiting infectious in groups most at risk [6], although a variety of social distancing policies have been explored in the literature. For instance, risk for Covid-19 and other influenza-like diseases vary widely by patient age, with increasing risk for hospitalization, poor prognosis, and death among older individuals [7] [8]. Prior work has explored the effect of different policies by considering isolation by age group [9] [10] [11], different modalities of social distancing [12], and different lengths of social distancing duration [9]. Several locations in the US have also varied their social distancing restrictions across time, with periods of stricter distancing and periods of re-opening businesses and social gathering places. These variations on policy invite the question of what the optimal social distancing policy would be, particularly if it can vary over time and be applied selectively to different demographics within the population.

However, to our knowledge, no prior work has examined social distancing policy duration by age group using optimization techniques. This is possibly due to the complexity of the optimization framework needed, as it would require integration of non-linear disease dynamics into the programming model. We do so in this work by solving a Markov decision process (MDP) model using simulation and numerical evaluation approaches. This is much more computationally intensive compared to more efficient value iteration or policy iteration algorithms [13] which are not feasible for this model setup. However, setting up the optimization framework in such a way allows us to include underlying disease dynamics in our optimization problem which are governed by a compartmental model of Covid-19 parameterized by empirical data and information from the medical literature. Critically, the compartmental model separately tracks the younger cohort (those aged under 55) and older cohort (those aged above 55) in the US. While age is not the only risk stratification relevant to Covid-19, we choose to focus on it here as a reasonable proxy for disease vulnerability; additionally, should a demographically heterogeneous social distancing policy be implemented, enforcement would be simpler if it varied only by age as opposed to unobserved biological risk factors (such as presence of comorbidities, etc., that also influence Covid-19 risk). We use an objective function that considers both social utility and health outcomes to determine when social distancing should occur provided a finite time horizon (e.g., when an effective treatment/vaccine is developed and disseminated). This optimization framework allows us to identify optimal social distancing policies that vary across age groups and time. While we do not consider a particular mechanism for achieving reduced social mixing (e.g., we are not specifically considering school closures, etc.), we hope that this general approach where contacts between certain age groups are reduced can provide valuable general insight into disease control policy.

We compare this demographically inhomogenous, time varying optimal policy with an optimal policy that cannot vary over age groups, as well as standard comparators where the whole population must either social distance or not over the entire time horizon. Examining these policies sheds insight into the effectiveness of social distancing by age group and the value of allowing the policy to change over time.

This work provides the following three main contributions: (1) we demonstrate how a compartmental model of disease transmission may be used in a Markov decision process to identify optimal disease control policy, despite the computational complexity; while the numerical example in this manuscript focuses on Covid-19, this approach is generalizable to many other infectious disease control problems where there is demographic variation. (2) We identify an optimal social distancing policy if demographic specific, time varying policies were being considered; (3) we quantify the benefit of such a policy compared to less flexible policies and explore the effect of parameter variation on our results.

## Methodology

### Overview

We construct a Markov decision process (MDP) model of a population meant to broadly approximate the US population to seek the optimal dynamic policy that maximizes the utility of a social distancing measure. In our model, in each epoch, a policy-maker can decide to implement social distancing or not; if so, individuals are assumed to mix only with household contacts.

To identify the number of infected individuals in each epoch, we employ a SEIR compartmental model of disease. In an SEIR model, the population is divided into four compartments: susceptible (S), Exposed (E), Infected (I) and Recovered (R). Because there is substantial evidence that there exists significant differences between young and old coronavirus patients, we model ‘young’ (below age 55) and ‘old’ (above age 55) cohorts separately in the SEIR model. The number of individuals in each group in the simulation roughly corresponds to the population proportions observed in the US. This allows us to capture and predict disease dynamics more precisely. This results in two joint SEIR models with eight compartments to describe the flow of population between different health states. We parameterize the model with data from the medical literature as well as empirical mixing pattern data. Some parameters, such as the overall transmission rate, were calculated – as in the case of the transmission rates between age groups, which used the next-generation method on information around *R*_0_ ranges for Covid-19.

Our objective function, the total social utility, includes benefits from social activity (which is lost when social distancing) and the negative effects due to disease infection (note that these, like the benefits of social activity, may not be financial). This disutility per patient might include average medical costs as well as reductions in the quality of life due to sickness and mortality and vary across age groups. The social benefit for individuals when not socially distancing includes the benefits of social activities, economic activity, and potential consumption. Because these costs and benefits include financial and psychological components, the exact value of these two parameters are difficult to obtain. We therefore pick values for these parameters that are not driven by data when simulating the model. These outputs, while perhaps not precise or realistic, still allow us to observe useful patterns in the optimal policy. We then additionally explore these values in sensitivity analysis to determine the robustness of these patterns.

We examine two policies. The first is where we assume policy makers can mandate social distancing or not in each time epoch, but is homogeneous across the population (the ‘homogeneous’ policy hereafter). The second policy additionally allows the policy to vary social distancing mandates across age groups as well as over time – because the policy varies across the population, we refer to this policy as the ‘inhomogeneous’ policy from now on. We compare the outcomes of these policies to scenarios where all individuals either do no social distancing or are required to social distance across the entire time horizon.

We assume that social distancing policies will stop being necessary after an effective vaccine is fully deployed. We therefore need to determine the optimal homogeneous and inhomogeneous social distancing policies assuming a finite time horizon, where the terminal time would be determined by when a vaccine is rolled out. We approximate this process by assuming the vaccine is fully rolled out quickly relative to the optimization time horizon, and assume the problem stops at time T (no gradual vaccine rollout and no immune state). We also separate this time horizon into discrete time epochs in which a single social distancing policy must be followed in each epoch; this is to prevent too-frequent policy changes, which would not be realistically implementable. For the purposes of our modeling example, we use a time horizon of 50 weeks and 5 epochs, resulting in epoch lengths of 10 weeks.

### Utility Function and Objective

Our goal is to maximize the total social utility before vaccines come out, which incorporates the social benefits earned from social activities when people go out and the disutility felt by those infected. In the homogeneous problem, the policy-maker considers using a social distancing policy in each epoch before vaccination occurs over the entire population. Suppose *x*(*t*) is a discrete decision variable that only takes values 0 or 1, where *x*(*t*) = 1 represents implementing social distancing and *x*(*t*) = 0 means it is not implemented. In this case, we model social distancing by modifying the contact matrix such that mixing between age groups is reduced to emulate reduced contacts outside the home (details given in the sections on transmission parameters below).

If the total population is *N*, and each person doing social activities can earn *s* benefit per unit time, then the momentary social benefit rate is *Ns*(1 − *x*(*t*)). We consider two age groups (young, below age 55, and old, age 55 and above). We denote *C*_*y*_ and *C*_*o*_ as the cost of coronavirus patients per unit time associated to the young and old age groups, respectively. By assuming that *I*_*y*_ (*t*) and *I*_*o*_ (*t*) are the number of young and old individuals infected, respectively, at time *t*, the total cost is given by *C*_*y*_*I*_*y*_(*t*)+*C*_*o*_*I*_*o*_(*t*). Thus, the momentary total utility for the homogeneous problem is:

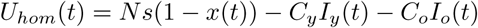

If *T* is the time when the vaccine comes out, we aim to find the optimal *x*(*t*), i.e., the optimal social distancing policy with respect to time, such that the total utility over the time horizon [0, *T*] is maximized. Mathematically, for the homogenous problem, our objective is:

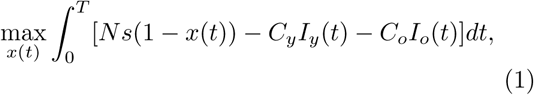

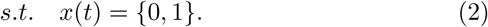

Note that this utility function gives 0 utility and 0 costs if everyone socially distances across the time horizon and no one becomes infected, providing a reasonable baseline for evaluation.

The utility function and decision variable above are for the homogeneous problem where a policy maker implements the same policy across the entire population. If one wishes to implement different polices across the young and old cohorts, then we can define similar decision variables, *x*_*y*_(*t*) and *x*_*o*_(*t*) for the young and old, respectively, and the momentary total utility would be:

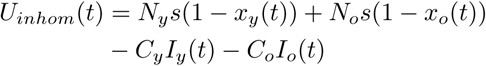

where *N*_*y*_ and *N*_*o*_ represent the young and old population. Thus, our objective becomes:

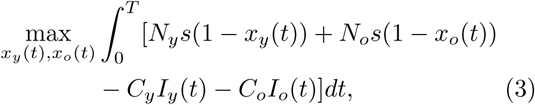

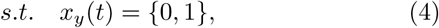

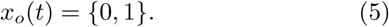

The intractability of the preceding optimization problem arises from the fact that *I*_*y*_(*t*) and *I*_*o*_(*t*) are captured by the susceptible-exposed-infected-recovered (SEIR) model, which is governed by a system of differential equations. The choice of policy at each moment before time *t* (i.e. the value of *x*(*u*) when *u* ∈ [0, *t*]) affects the transmission rate in the SEIR model so that implemented policies can influence *I*_*y*_(*t*) and *I*_*o*_(*t*) in complex ways.

### Markov Decision Process (MDP) Formulation

To overcome this difficulty, we separate the time horizon into several equal time slots and require that the same policy be implemented over the whole time slot. This is more likely to match the real world because changing policies frequently will be inconvenient and difficult to implement.

Suppose we separate time horizon into *n* equal time slots where the *i*_*th*_ time slot is represented by [*T*_*i*_, *T*_*i*+1_] where *i* ∈ {1, 2, …, *n*} and *T*_1_ = 0, *T*_*n*+1_ = *T*. At the beginning of time slot *i*, the policy maker decides which policies to implement (enforce or not enforce social distancing), i.e. select the value of decision variable *x*_*i*_ from the set {0, 1}. The initial state of *I*_*y*_ and *I*_*o*_ at time slot *i* is determined by the initial state of *I*_*y*_ and *I*_*o*_ at time slot *i* − 1 and *x*_*i*_, so we can reformulate our previous optimization problems (1) and (2) into a Markov decision process (MDP) problem. MDPs are widely used across multiple disciplines for sequential decision-making problems, where there are repeated opportunities to make decisions and the optimal action now depends on future actions. MDPs have been widely used in prior literature in healthcare policy: for example, Alagoz et al. formulated a MDP model for optimal timing of liver transplantation [14], and Denton et al. constructed a MDP model to optimize selection of patients with type 2 diabetes for statin therapy [15].

In our MDP model, states include the size of the susceptible population (S), the number in the exposed state (E), the number of infected individuals (I), and the number of people in the recovered state (R). The transition function is governed by the age-stratified SEIR model and the selected action. In the homogeneous problem, where the social distancing policy can vary over time but not the population, the action space contains only two elements: implementing social distancing and not implementing social distancing. In the inhomogeneous problem, where decision makers can implement different actions for the young and old cohorts, the action space will contain four elements. For simplicity, denote 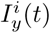 to be the number of young individuals infected at time *t* of the *i*_*th*_ time slot, and 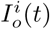 for the old individuals. Thus, our homogeneous problem MDP formulation is:

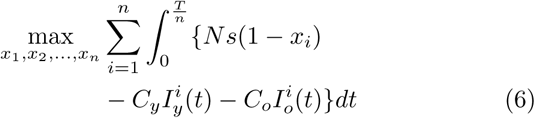

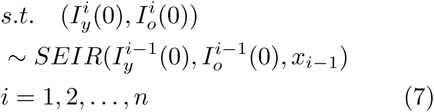

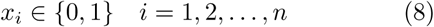

Since the length of each epoch is 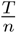, we treat 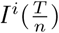 as the end state of the epoch i, and the initial state in each epoch is equivalent to the end state of the last epoch: 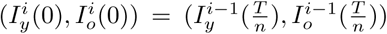 The inhomogenous problem formulation is similar, except that we use *U*_*inhom*_(*t*) for the utility function to capture social benefits separately for young and old, as they may have different social distancing actions.

Given the relatively small number of epochs in our problem, we can solve both MDPs through simulation and numerical evaluation. For each possible social distancing policy, we first calculate the total utility by running the SEIR model under that sequence of social distancing policy in each epoch. We then compare the total utility across all 64 possible social distancing policies to determine which one is best. We repeat this procedure to identify the optimal inhomogeneous social distancing policy, where there are 1024 possible policies.

### Infection Cost and Social Distancing Benefit Parameters

Since the costs of social distancing (financial, social, and psychological) are difficult to quantify, we assign the social activity value of benefit per capita to have a value of 1 (this is the value that is lost when social distancing policies are implemented). The relative health and mortality costs are similarly difficult to quantify, but we may imagine them to be much larger (as individuals may die or have long term health effects from infection). We therefore assign the costs per patient to be 100 and 200, for young and old infected cases, for each epoch an individual is infected. In sensitivity analysis, we modify patient health costs to vary between 25 and 800 for young and between 50 and 1600 for old, respectively, which we believe is a sufficiently large range to capture reasonable variation in outcomes.

### Age-stratified SEIR model

In order to tackle our MDP problem, we build an age-stratified SEIR model wth two age groups to simultaneously describe the population dynamics for young and old. In this model, we separate the population into young and old groups and assume cross-infection happens in both groups. Thus, the age-stratified SEIR model is expressed as:

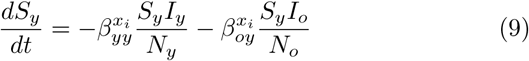

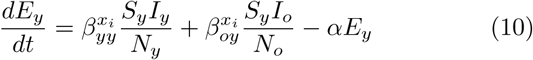

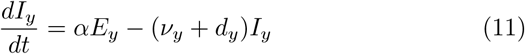

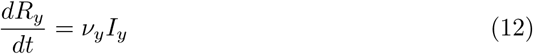

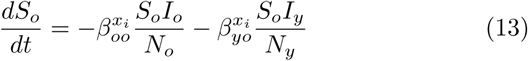

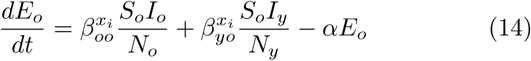

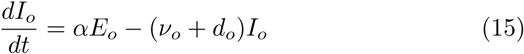

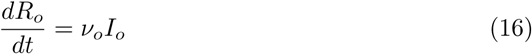

Here, subscripts *o* and *y* denote old and young respectively, and 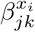 represents the transmission rate at which individuals in age group *k* become infected by age group *j* under policy *x*_*i*_. *α* represents the rate of disease activation (from latent to active stage), *ν* is the clearance rate, and *d* the death rate. Figure 1 depicts each compartment’s inflow rates and outflow rates in the SEIR model. We assume higher mortality rates for old patients (*d*_*o*_ ≥*d*_*y*_) to better capture the poorer health outcomes experienced by this particularly high-risk group.

**Figure 1:**
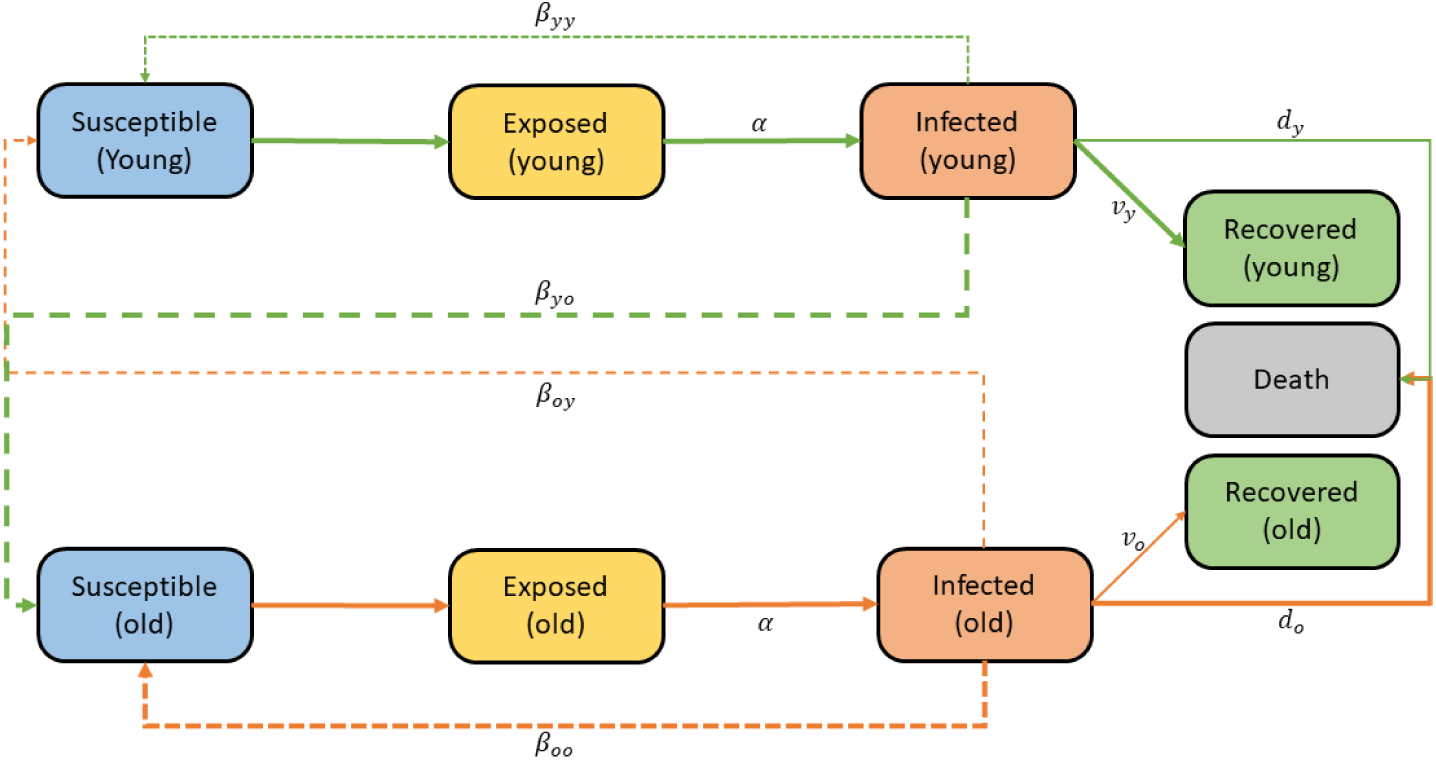
Model Schematic

### Estimation of SEIR Model Parameter Values

The accurate estimation of parameter values is crucial to complete the age-stratified SEIR model and solve the MDP problem. However, some parameters, like transmission rates, can not be found from the literature directly, or are known only with great uncertainty. Others show considerable variation, as there exists heterogeneity across different settings in the US. Additionally, this SEIR model is age-specific, and there may not be research examining values across different age groups. In the following sections, we will introduce how we estimate model parameters and how we address these challenges.

#### Incubation Period

The incubation period is the time from exposure to the causative agent until the first symptoms develop. We assume there is no significant difference in incubation periods between young and old. Hence, the parameter 1*/α*, representing the incubation period in the age-stratified SEIR model, is the same for both age groups. Based on the findings of Li, Guan, et al., we set 1*/α* to be 5.2 days [16].

#### Mortality Rate

The mortality rate is a measure of total deaths per unit time in a particular population, scaled to this population’s size. Thus, the mortality is equal to the product of the case fatality rate and the inverse of the duration from symptom onset to death. The study from Wu and McGoogan shows that the general case fatality rate in China is 2.3%, but it jumps to 8% for those aged between 70-79, and 14.8% in those aged between 80-89 [17]. Thus, in our age-stratified model, we consider the heterogeneity of mortality rates in different age groups.

The CDC website provides a report of demographic trends of Covid-19 cases and deaths, from which we calculate mortality values (shown in Appendix Table A.1 and A.2 [18]). We use age 55 as the cutoff between young and old groups in the age-stratified SEIR model. Thus, the deaths and confirmed cases in the young group is 14,815 and 3,435,721, respectively, and is 128,142 and 1,407,712, for the old cohort. As a result, the case fatality rate for the young group is 0.4% and10% for the old group. For the number of days from symptom onset to death, Wang et al. found that the average time from the first symptom to death is about 14 days, and this value will decrease to 11.5 days in ages older than 70 and increase to 20 days in age younger than 70 [19]. To use these observations for our age stratification, we set the average time from symptom onset to death among young and old to be 25 and 13 days, respectively, which matches the trend that older patients die faster than younger patients. Therefore, the mortality rates for young and old are *d*_*y*_ = 0.00016, *d*_*o*_ = 0.0077. However, we recognize that there is considerable variation in mortality values across different geographical regions, health systems, and demographic groups, and we therefore perform a sensitivity analysis around these parameters. In these sensitivity analyses, we vary the mortality rate from 0.0001 to 0.00025 for the young cohort and 0.006 to 0.009 for the old cohort to observe changes to model outcomes.

#### Clearance rate

The clearance rate should be equal to the product of the proportion of recovered individuals and the inverse of days until recovery. Given the case fatality rate information above, the recovery rate among young and old are 99.6% and 90%, respectively. Pan et al. found that the average recovery time since the initial onset of symptoms is 10 days [20]. However, to the best of our knowledge, there are no data tracking the recovery time grouped by age, so we use 10 days as the average days until recovery for both age groups. The clearance rates for young and old are then *ν*_*y*_ = 0.1 and *ν*_*o*_ = 0.09.

In sensitivity analysis, we vary the clearance rate. The observed data reveals that the range of days until recovery is roughly between 5 to 16 days [21], so the ranges of clearance rate for young and old are 0.06 to 0.199 and 0.056 to 0.18, respectively. We increase clearance rates by 0.05 steps in both groups, with all other parameters kept constant (note that this means that the resultant *R*_0_ varies across sensitivity scenarios). Since young people usually have stronger immune systems, we set clearance rate for the younger cohort such that they are always higher than that for old individuals.

#### Transmission Input Data

The values for the four transmission parameters *β*_*yy*_, *β*_*yo*_, *β*_*oy*_ and *β*_*oo*_ were informed by constructing population-level contact mixing matrices. Following the approach taken by Vardavas et al. [22], we use two sources to derive these values, one for our base case analysis and the other as a sensitivity analysis scenario.

For our base-case analysis, we use available data provided by the Network Dynamics and Simulation Science Laboratory (NDSSL) at Virginia Polytechnic Institute and State University that represent a synthetic population of Portland Oregon [23]. The NDSSL data for Portland, Oregon, provides an instance of a time-varying social contact network for a normative workday, derived from daily activities. The data were created from an urban transportation agent-based model (ABM), which simulated individuals’ daily movements across locations in Portland, Oregon. The data are synthetic and represent only Portland on a usual workday, and so might not be indicative of the entire United States averaged across all days.

The data provides an edge-list that specifies which nodes or vertices representing individuals are connected. The edge-list data further contains information regarding each individual’s activity or purpose during the interaction, and an edge-weight is given by the duration, in seconds, of the interaction. Examples of these activities include home, work, leisure, school, and commerce. More than 90% of all edges in the edge-list date are assortative in the two connected individuals’ purpose – the vast majority of edges describe interactions where both individuals engaged in the same purpose (e.g., Home-Home, and Work-Work interactions). We thus categorized each edge with a single purpose describing the nature of the interaction. For example, Home-Home edges were categorized as Home interactions. We also consolidated the many purposes into six main categories: Home, Work, School, Commerce, Leisure, and Other. All edges with disassortative purposes of the two connected individuals were categorized as an Other. The edge-list data was used to generate the conditional mixing matrices that give the relative proportion of contacts of an individual belonging to a given population stratum (i.e., age group) with the other strata for the six purposes. Moreover, from the edge-list, we extracted the six coefficients representing each of the six mixing matrices’ relative matrix-weights. These matrix-weights sums to 100% and were calculated by summing the edge-weights for each of the six interaction categories. Since the matrix-weights are based on edge-weights rather than on the tally of each edge type, they represent each type of interaction’s overall strength in terms of aggregate duration rather than on aggregated interaction-tick frequency. In this study, we used these six mixing matrices and consolidated them into two mixing matrices, one describing home-level mixing and the second describing all other mixing patterns. Overall matrix-weights were calculated and associated with the two mixing matrices. To model social distancing, we scale down the importance of the matrix-weight associated with all other interactions while keeping the matrix-weight associated with home mixing unaltered. After scaling, the sum of the two matrix-weights is smaller than 100%. This data was used for the base case analysis and is shown in Table 1.

**Table 1:**
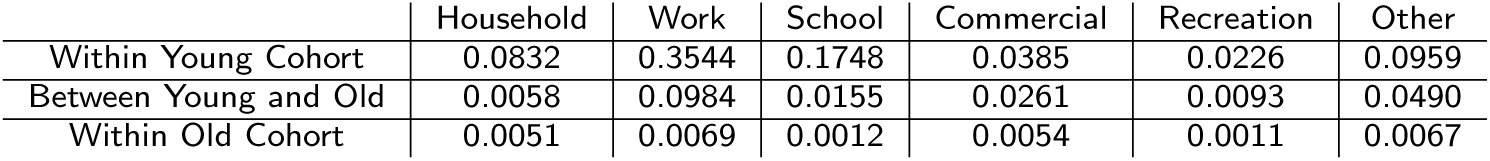
Proportion of total contact calibrated by age and locations in the base case (calculated from NDSSL data)

The second data source [24] was used for our sensitivity analysis. It is based on self-reported survey data of unique contacts over the course of one day in eight European countries, listing the age, sex, and location of the contact [25]. This data has been extrapolated to create mixing matrices for 152 countries, including the United States [24]. Similar to our procedure on the NDSSL data, the location was used to create age-specific mixing matrices associated with interactions that occur at the categorical groups Home and Other. As before, associated with each of the two mixing matrices, we computed their relative weights, which add to 100% under pre-pandemic status-quo conditions. Social distancing is modeled by scaling down the matrix-weight, multiplying the mixing matrix associated with interactions belonging to the category Other.

The mixing matrices produced by these two sources are quite different, and we therefore use both to examine the effect of variation in transmission on our model outcomes. We chose to use the mixing matrices generated from the NDSSL data for our base case analysis because the NDSSL was generated from detained US data for Portland, OR. The second data source [24] was used for our sensitivity analysis because the data was originally generated for European countries and later transformed to be representative of the US.

However, this process only provides the relative values of *β*_*yy*_, *β*_*yo*_, *β*_*oy*_ and *β*_*oo*_; we still do not know their actual values. To determine that, we calibrate these values using *R*_0_. We describe this process in the next section.

### Transmission Rate Calibration

We apply the next-generation method to calibrate transmission rates, which derive basic reproduction numbers (usually denoted as *R*_0_) from a compartmental model of infectious disease. Based on the age-stratified SEIR model and partial estimated parameter values, transmission rates could be calibrated using the nextgeneration method to generate an *R*_0_ value of 2.4 [26].

We have eight independent transmission rates (as we have two age groups and two social distancing settings). Thus, we explore the relationship of each transmission rate using its definition: the transmission rate is equal to the product of the probability of infection given a contact between a susceptible and infected individual (*ρ*) and the average contact rate between two groups (*c*). Define 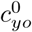 to be the average contact rate between young to old under policy *x*_*i*_ = 0, then 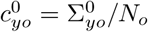 where 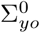 is the number of total contacts between young and old when *x*_*i*_ = 0 and *N*_*o*_ is the number of old individuals in the population. If we do not consider the heterogeneity of *ρ* by age, then the ratio of any two transmission rates is equal to the ratio of corresponding average contact rates. For example,

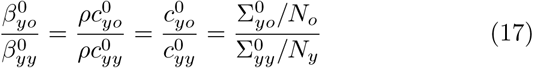

Since the number in the young population and old population is known, the ratio between other transmission rates and the base transmission rate 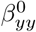 can be calculated from equation 17 combined with Table 1. For instance,

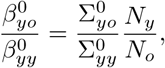

where 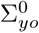 is equal to the sum of second row of Table 1 and 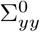 is equal to the sum of first row of Table 1.

Then,

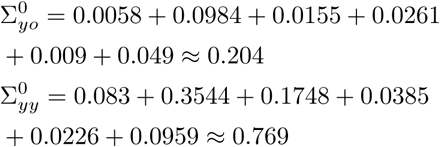

Then we can obtain

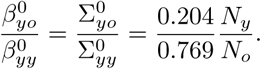

Similarly, we can calculate other ratios of transmission rates when social distancing is not in effect, which are organized in Table 2.

**Table 2:**
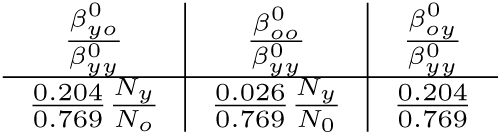
Ratios between partial transmission rates and base transmission rate.

If 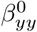 is known, then 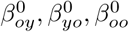 can be obtained by Table 2. Diekmann et cl. illustrated that one can determine *R*_0_ from compartmental models by decomposing its Jacobian matrix into *V* +*F*, where *V* roughly represents the production of new infections, and *F* represents transitions, describing the change of state. *R*_0_ is then equal to the largest eigenvalue of −*V F* ^−1^ [27]. Using the compartmental model, we derive

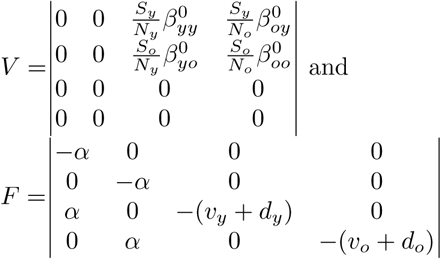

where *S*_*y*_ and *S*_*o*_ are the number of young and old susceptible individuals corresponding to the time when *R*_0_ = 2.4, the reproductive number when no social distancing policy was in effect. Replacing Initial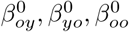 by 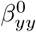 and setting *N*_*y*_, *N*_*o*_ as shown in Table 5, based on Table 2, then we can calculate *R*_0_ given any given 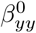. This relationship is shown in the plot in Appendix Figure A.1.

### Social Distancing Assumptions

To model social distancing policies, we assume contact only occurs within households. This means that we set all other contact modalities besides Home to be 0 in our transmission data for this calculation. If the old do social distancing but the young do not, we assume the contacts between the young and the old also only occur in the household. For example, if we want to calculate 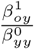 we first use an equation similar to equation 17:

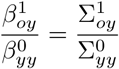

To calculate 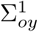 when social distancing, we need only sum the contacts between young and old that occurs in the household. Hence,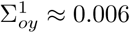, and

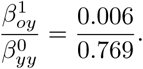

We can calculate other transmission rates under social distancing in a similar way. These are provided in Table 3.

**Table 3:**
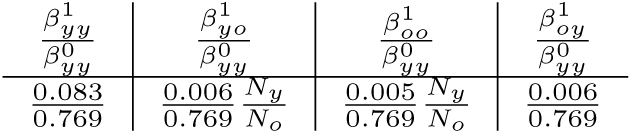
Ratios between partial transmission rates and base transmission rate.

Using Appendix Figure A.1, the estimated 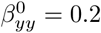 if *R*_0_ is 2.4. Then, we use Tables 2 and 3 to calculate all transmission rates, shown in Table 4.

**Table 4:**
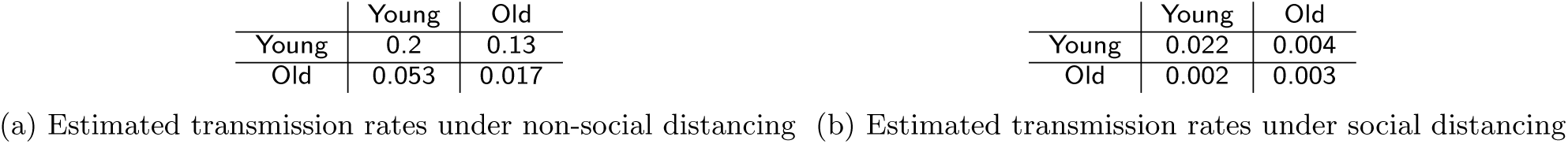
Estimated transmission rates.

### Initial Conditions

We start the simulation with 30 individuals infected, 120 individuals exposed, no recovered individuals, and 300 million individuals total, which were the figures near the beginning of the epidemic in the US [28]. The initial value of *I*, 30, is the sum of *I*_*y*_ and *I*_*o*_. The exact value of *I*_*y*_ and *I*_*o*_ depends on the ratio of individuals in the young and old groups. Since we stratify the population using age 55 as a cutoff, the proportion of the young population is 0.71% [29], so the ratio of young and old population is 0.71*/*0.29 =2.45. Thus, the initial values of *I*_*y*_ and *I*_*o*_ are 21 and 9 respectively.

The initial value of *E* is set as four times of *I* [28]. Using a similar rationale, *E*_*y*_ and *E*_*o*_ are then 84 and 36, respectively. Table 5 lists all base case parameter values.

**Table 5:** Parameter values of base case.

| Notation | Description | Value | Unit |
| --- | --- | --- | --- |
| $N$ | Total population | 300,000,000 | No. of People |
| $s$ | Social benefits per capita | 1 | $(\text{No. of People} \times \text{Day})^{-1}$ |
| $D_y$ | The cost per capita for young people | 100 | $(\text{No. of People} \times \text{Day})^{-1}$ |
| $D_o$ | The cost per capita for old people | 200 | $(\text{No. of People} \times \text{Day})^{-1}$ |
| $S_y(t=0)$ | The number of susceptible young people at time 0 | 213,000,000 | No. of People |
| $S_o(t=0)$ | The number of susceptible old people at time 0 | 87,000,000 | No. of People |
| $E_y(t=0)$ | The number of exposed young people at time 0 | 84 | No. of People |
| $E_o(t=0)$ | The number of exposed old people at time 0 | 36 | No. of People |
| $I_y(t=0)$ | The number of infected young people at time 0 | 21 | No. of People |
| $I_o(t=0)$ | The number of infected old people at time 0 | 9 | No. of People |
| $R_y(t=0)$ | The number of recovered young people at time 0 | 0 | No. of People |
| $R_o(t=0)$ | The number of recovered old people at time 0 | 0 | No. of People |
| $\nu_y$ | Clearance rate of young patients | 0.1 | $\text{Day}^{-1}$ |
| $\nu_o$ | Clearance rate of old patients | 0.09 | $\text{Day}^{-1}$ |
| $d_y$ | Death rate of young patients | 0.00016 | $\text{Day}^{-1}$ |
| $d_o$ | Death rate of old patients | 0.0077 | $\text{Day}^{-1}$ |
| $\alpha$ | Incubating rate from exposed state to infected state | 0.192 | $\text{Day}^{-1}$ |

### Sensitivity Analysis

This analysis depends on many estimated parameter values. However, these values are uncertain for a variety of reasons – heterogeneity over the population, measurement error, lack of evidence, etc. We therefore perform sensitivity analyses to determine the robustness of the identified optimal policy. In particular, we examine sensitivity scenarios on the utility parameters, transmission rate, clearance rate, and mortality rate. Parameter ranges for these analyses are given in the Appendix (Additional Sensitivity Analyses section).

## Results

### No Social Distancing

Using the policy where no social distancing occurs, a large part of the population, both young and old, become infected. The social utility for young individuals over the time horizon is −1.52 per capita and −3.20 per capita for old individuals. This results in a total of 1.26 sick-weeks per capita among the young population over the time horizon and 1.05 sick-weeks per capita among the old population (see Figure A.3a). In the end, there would be 5.2M deaths and 245.5M people in the recovered state. Outcomes from all policies are summarized in Table 6.

**Table 6:**
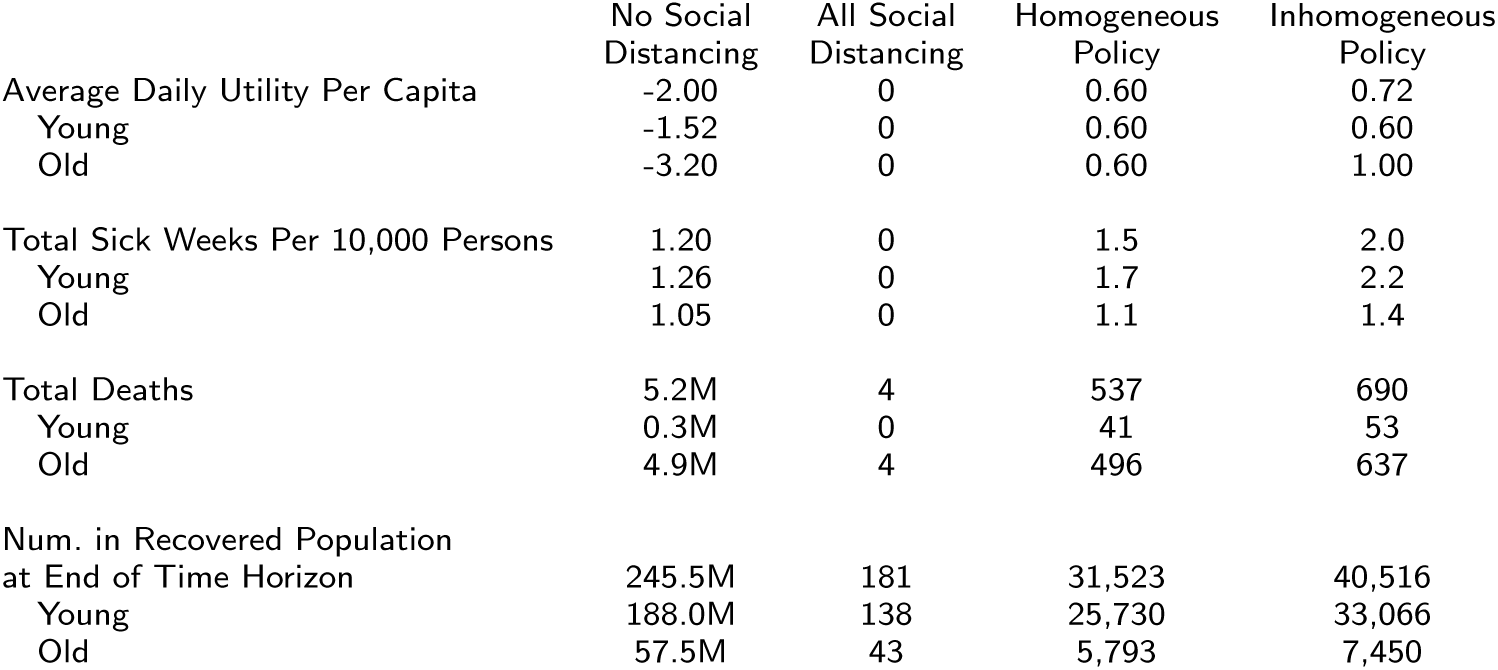
Outcomes across all policies.

### Social Distancing for the Entire Time Horizon

By contrast, if all individuals are socially distanced for the entire time horizon, most of the population remains in a susceptible state, with few individuals contracting illness. The number of individuals infected falls to almost 0 after 100 days’ social distancing, greatly reducing the total number of infected over the time horizon. There would be almost 0 sick-weeks per capita among the young and old populations over the time horizon. As the number of infected is drastically reduced, at the end of the time horizon, virtually no one would be in the recovered state: 138 and 43 individuals would be in the recovered state for young and old populations, respectively (see Figure A.3b), compared to 188.0M and 57.5M in the no-social distancing scenario. This also greatly reduces mortality over the time horizon, with only 4 deaths.

These health gains outweigh the social cost of distancing using our base case utility parameters, with an average social utility of 0 among the young population and 0 among the old population, an increase of 1.52 and 3.20 from the no-social distancing case. Hence, in terms of social benefit and health gains, the all-social distancing case outperforms no-social distancing case. However, note that the young population bears the brunt of the disease burden in both cases.

### MDP Results: Homogeneous Policy

While all or none social distancing policies are natural first options, additional social utility and better health outcomes may be garnered if the policy can change over time. We examine this scenario using the MDP solution to the homogeneous policy problem, where the optimal policy is to socially distance only in the first and second epochs (out of five total epochs over the time horizon; each epoch is 10 weeks).

The disease will be controlled well under the optimal homogeneous policy where about 22,430 individuals are infected at the end of simulation and a very small number of people become infected during the first 300 days (shown in Appendix Figure A.2a). With this policy, we find that social utility is higher than both extreme distancing scenarios. The total social utility would be 0.60 per capita, with 0.60 per capita accrued to the young population and 0.60 utility per capita to the old population, respectively. This is in contrast to the preceding all or none scenarios, which had lower overall utility for both groups. This improvement is in part due to the shorter duration of social distancing as well as the maintenance of better health outcomes than the no-social distancing case.

In terms of health outcomes, this optimal policy would result in a total of 0.0002 sick-weeks per capita among the young population and 0.0001 sick-weeks per capita sick-weeks among old population, respectively (see Figure A.3c), an increase of of 0.0002 sick-weeks per capita and 0.0001 sick-weeks per capita from the all-social distance policy. Additionally, 537 people would die and 31,524 people end up in the recovered state.

These epidemiological results are quite sensitive to when social distancing starts. If we run the simulation and delay the optimal social distancing policy by one epoch (socially distance only in the second and third epochs), the peak number of individuals infected reach more than 8,000 people (shown in Appendix Figure A.2b). This comparison reveals that only a one-epoch delay in social distancing will result in an outbreak.

### MDP Results: Inhomogeneous Policy

We additionally consider the case where we allow for additional flexibility in policy where there can be different social distancing policies for the young and old (an inhomogeneous, time-varying policy). In this case, the optimal policy is for the young to socially distance in the first and second epoch while the old never socially-distance. The epidemiological change over time is shown in Appendix Figure A.3d. Compared with a homogeneous optimal policy, the inhomogeneous policy would result in higher total utilities over the whole population, but not within the young population. Health outcomes also worsen, as less social distancing occurs. The total utility per capita increases by 0.12 compared to the optimal homogeneous policy, while the utility per capita among the young population almost does not change. The overall utility per capita increase is due to the improvement among old population, which increases by 0.40 compared to the homogeneous policy(see Table 6).

The inhomogeneous policy results in worse health outcomes than the homogeneous policy. The per capita sick-weeks of the young and old population nearly does not change(see Table 6). However, less total social distancing must result in more infections, which translates to 690 deaths in the inhomoegenous policy, 153 more deaths than with the homogeneous policy. The inhomogeneous policy also results in 40,516 people in the recovered state, which is 8,992 more than recovered people as under the homogeneous policy.

Overall, however, these health decrements are not nearly as drastic as the ones seen when compared to the no social distancing policy, although old individuals never social distance. This is for a few reasons. One is that the transmission rate between socially-distancing young and non-socially-distancing old is same as in the case when both the young and the old socially distance, because in both cases the two groups would only contact each other in the home due to our assumptions about social distancing. In addition, the transmission rate between old individuals under social distancing and no-social distancing policies are 0.003 and 0.017 (see Table 4), respectively. Both of these values are relatively small, which means old people do not contact each other much in either case. Lastly, old people occupy a small part of the total population, which mitigates the risk of infection from this subgroup.

### Results: Summary

In summary, social distancing can indeed mitigate the negative health outcomes of pandemics and preserve utility, even when within-household contacts continue. Unsurprisingly, increased health benefits through decreased transmission and infection have the potential to outweigh lost social utility. While this depends heavily on the magnitude of utility from social contacts, which we explore in sensitivity analysis, our base case analysis shows no social distancing can generate far worse outcomes on both health and social utility fronts.

This analysis also demonstrates that using an inhomogeneous policy that varies across time and by demographic group can preserve utility with only small compromises to health outcomes. Surprisingly, in the optimal inhomogeneous policy, we find that young individuals have nearly the same social distancing policy as in an optimal homogeneous policy. Old individuals, by contrast, do not stop social activities at all. Our transmission input values explain this unexpected outcome. Our data shows a low transmission rate among old individuals – even when they do not socially distance, they are relatively much less likely to contact others than younger individuals. The majority of young-old contacts occur in the household, which is unchanged due to social distancing. While we vary these transmission patterns in sensitivity analyses described below, it is interesting to note that reasonable contact patterns can generate unexpected outcomes.

As mentioned, these results may depend heavily on transmission rates, mortality, and social utility costs. Since great uncertainty remains in what these values may be, we perform sensitivity analyses to examine the effect variation would have on our results.

### Sensitivity Analysis Results

We perform sensitivity analyses on a variety of parameters. However, the utility function benefits and costs are the most uncertain and they also lead to qualitatively different results, so we summarize the results from the other sensitivity analyses here (full sensitivity analysis in the Appendix) and present utility parameter sensitivity results in the main text.

In sensitivity analyses around utility and costs, we find that varying the costs for young and old patients, respectively, between 25-800 and 50-1600 did not change the optimal policy. It was still optimal to social distance the first and second epochs (both groups for the homogeneous policy; only young for the inhomogeneous policy).

In clearance rate sensitivity analyses, the length of social distancing needed decreases as clearance rate increases. Both homogenous and inhomogeous social distancing policies are sensitive to clearance rate variation over the range we examined, although social distancing epochs remained concentrated in the earlier half of the time horizon. With sufficiently high clearance rates, social distancing may not be necessary to preserve utility, although health outcomes may deteriorate slightly in this scenario.

In the transmission rate sensitivity analysis, we use the mixing contact matrix from [24]. We find the optimal policy is to socially distance in the second and third epoch (all individuals in the homogeneous policy; only young indivdiuals in the inhomogeneous policy). Due to having larger transmission rates and the changed optimal policy for the mixing matrix, the health outcomes worsen in this scenario compared to the base case. As in the base case, the inhomogeneous policy outperforms the homogeneous policy in utility outcomes.

Another interesting observation concerns the optimal time to start social distancing. In both the homogeneous and inhomogeneous policy under this alternative contact matrix, the optimal time to start social distancing is not at the very beginning of the epidemic – socially distancing immediately may not always the best choice. Intuitively, this occurs if the epidemic has not yet grown to sufficient size to warrant the lost utility from social distancing. These optimal policies also do not recommend social distancing up until the time of a vaccine roll out – instead, they stop a few epochs before, even in the homogeneous policy where both young and old social distance. This is because, at some point in time, the infection does not have sufficient time to spread widely enough to outweigh the social benefits in the time remaining before a vaccine is found. The exact timing of optimal initialization of social distancing will depend on the disease dynamics and the magnitude of utility loss due to distancing.

We find that the optimal policies for both inhomogeneous and homogeneous problems are not sensitive to the mortality rate. As seen in the base case results, the inhomogeneous policy outperforms the homogeneous policy in terms of utility. Total sick weeks decrease and total deaths increase as mortality rate increases, as expected.

One takeaway of these analyses is that a limited amount of social distancing near the beginning of the epidemic can greatly reduce sick-weeks and total deaths. An MDP framework is a valuable tool to identify when this social distancing should occur as well as which populations to socially distance.

## Conclusions

In this work, we examine the optimal social distancing policy to maximize total social utility. We examine both social utility and health outcomes in the resultant policies. We do so by designing a novel MDP model that uses an age-stratified SEIR compartmental model to describe disease dynamics. This allows us to determine which groups (young or old) should be socially distancing at what times. While data on Covid-19 is continually being updated, we used information from the medical literature and empirical contact pattern data to calculate reasonable parameters for the model.

Using this framework, we identified both the optimal time-varying and time-homogeneous social distancing policies. The time inhomogeneous policy generated the most total utility, due to its increased flexibility. Both policies had young individuals social distancing in the first and second epochs; however, the policies varied much more widely for older individuals – in the time inhomogeneous policy, the optimal policy did not have older individuals social distance at all. This difference in policy results in higher social utility for old individuals and only slightly worse health outcomes for all individuals (in terms of mortality and individuals exposed to infection). We find that this result is primarily driven by the mixing patterns used in our model, which were derived from NDSSL [23]. This data indicated that older individuals did not do much socializing (and therefore transmit disease) even when no social distancing policy was in effect for them. In our sensitivity analysis, we found that these patterns were generally robust to changes in input values, and, critically, cost and benefit assumptions.

Our results demonstrate the value of using an MDP framework to examine optimal disease control policies which can change over time, particularly in the context of heterogeneous risk groups within the population. We were able to identify social distancing policies where infectious cases were virtually eliminated by imposing a high cost of infection; in these cases, the number of social distancing epochs remained small, and the policy varied when it was allowed to vary social distancing actions across demographics. We stratified the model in this analysis on age as older groups have been documented to have higher mortality and morbidity risks for Covid-19; however, this framework is applicable to other diseases as well, where it may be appropriate to differentiate risk groups along different characteristics. This would still be within the capabilities of this model framework, as it can handle any disease dynamics able to be captured by a compartmental model.

There are several limitations to this work which we would like to acknowledge here. First, there are several highly uncertain input parameters. In particular, the social benefits and cost values in our objective function are not driven by data; these values are difficult to measure, as they include psychological harms and stressors of social distancing, and empirical data is lacking. Instead, we perform sensitivity analysis around these values to show how our results would change under different values. Our results illustrate how identifying the pattern of social distancing using a MDP framework can still identify useful patterns for optimal policy design despite highly uncertain model inputs.

Secondly, the model structure (an SEIR compartmenal model) is a highly abstract description of true Covid-19 dynamics. In reality, transmission, recovery, and mortality, due to Covid-19 are dependent on a variety of individual characteristics not captured here – co-morbidities, social economic status, geographic location, poverty level, access to healthcare, etc. However, the simplicity of our model allows for more tractable simulation, and we argue that the results still provide general insights into optimal disease control policy.

Despite these limitations, this work demonstrates that, even across a wide range of social benefit values and health costs, social distancing can be a valuable tool for reducing infection. However, demographic mixing patterns and disease dynamics should be considered when designing such policies, as limiting social distancing restrictions to groups that mix more (such as young individuals in this model) can be an effective method of reducing the impact of social distancing policies for small changes in health outcomes. Moreover, social distancing policies should vary over time; starting too late may unnecessarily reduce social benefits without much reduction in infection.

## Data Availability

All data produced in the present study are available upon reasonable request to the authors.

https://journals.plos.org/ploscompbiol/article?id=10.1371/journal.pcbi.1005697

https://www.vtcrc.com/tenant-stories/virginia-bioinformatics-institute/

## Acknowledgements

Not applicable.

## Abbreviations

MDP: Markov Decision Process

homog: homogeneous

inhomog: inhomogeneous

## Availability of data and materials

The datasets used and analysed during the current study are available from the corresponding author on reasonable request.

## Ethics approval and consent to participate

No ethics approval was necessary.

## Consent for publication

Not applicable.

## Authors’ contributions

PD, RV, SN, SCS conceived the study; PD, SCS contributed to the problem formulation and analysis; PD, SCS, RV, SN wrote and revised the manuscript; PD, RV, SCS developed methodology and created the models; PD performed the formal analysis and implemented the computer code.

## Appendix

### Additional SEIR Model Parameters

In this section we present additional details on the SEIR model parameters.

#### Transmission

**Figure A.1:**
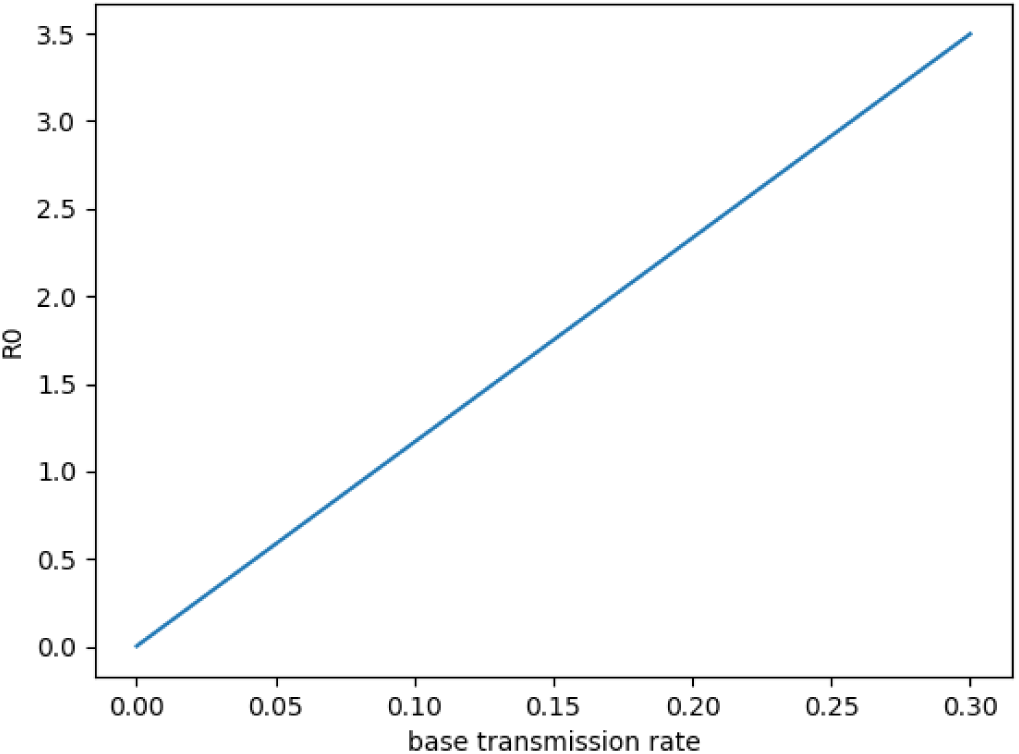
*R*_0_ & Base transmission rate.

In Figure A.1, we show the relationship between the base transmission rate (as described in the manuscript) and the corresponding basic reproduction number *R*_0_. This plot is generated by using next generation method where we input the base transmission rate as a variable and obtain *R*_0_. Since the estimated *R*_0_ is 2.4, we use a base transmission rate of 0.2 in our analysis.

#### Demographic Trends of Covid-19 Cases and Deaths

Appendix Table A.1 shows the confirmed number of Covid-19 cases and Appendix Table A.2, below, shows the number of Covid-19 deaths, by age group, in the US by August 2020; this information is taken from CDC website [18]. We use this information in the model to determine the mortality rate of different age groups. We do this by summing over all age groups from 0-55 to find the number of infected individuals and deaths under age 55; a similar procedure is done to determine values for the old cohort.

**Table A.1:** Confirmed cases by age.

| Age Group | 0 - 4 | 5 - 17 | 18 - 29 | 30 - 39 | 40 - 49 | 50 - 64 | 65 - 74 | 75 - 84 | 85+ |
| --- | --- | --- | --- | --- | --- | --- | --- | --- | --- |
| Count | 85,864 | 319,990 | 1,133,167 | 814,387 | 746,949 | 1,006,092 | 367,878 | 210,868 | 158,238 |

**Table A.2:** Deaths by age group.

| Age Group | 0 - 4 | 5 - 17 | 18 - 29 | 30 - 39 | 40 - 49 | 50 - 64 | 65 - 74 | 75 - 84 | 85+ |
| --- | --- | --- | --- | --- | --- | --- | --- | --- | --- |
| Count | 34 | 55 | 749 | 1,913 | 4,609 | 22,366 | 30,121 | 37,781 | 45,329 |

### Additional Results Graphs

We show the number of infected individuals over time under different social distancing policies in a series of plots. We examine what occur if the social distancing policy is implemented one epoch later in Figure A.2b than in Figure A.2a, which shows the optimal policy (social distance in epoch 1 and 2). Figure A.2a, using the optimal policy, shows that the number of infected increases at the end of the time horizon. Conversely, in Figure A.2b, there is a peak at about day 80. In this case, social distancing begins at the second epoch starting at day 70, so the disease will be suppressed when just as it is about to spread widely.

In addition, both Figures A.2a and A.2b show a rise at the end of time horizon, which occurs due to our objective to maximize social utility within the time horizon – after some time, suppressing the disease through social distancing is no longer worth the lost social utility given the limited amount of time until the end of the time horizon (when everyone is assumed to become immune to the disease)..

**Figure A.2:**
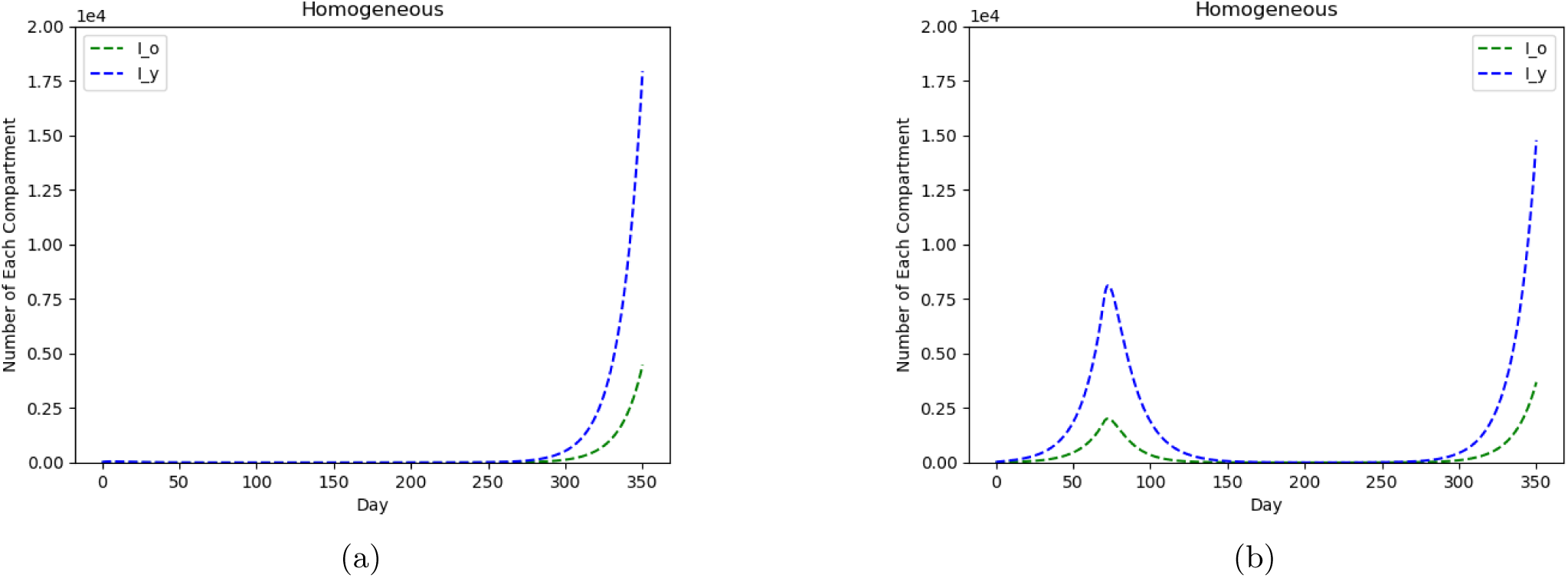
Sensitivity of infected population patterns to timing of homogeneous social distancing policy.

Figure A.3 depicts the epidemiological outcomes with each social distancing policy. Note that the y-axes scales are quite different to allow for readability.

**Figure A.3:**
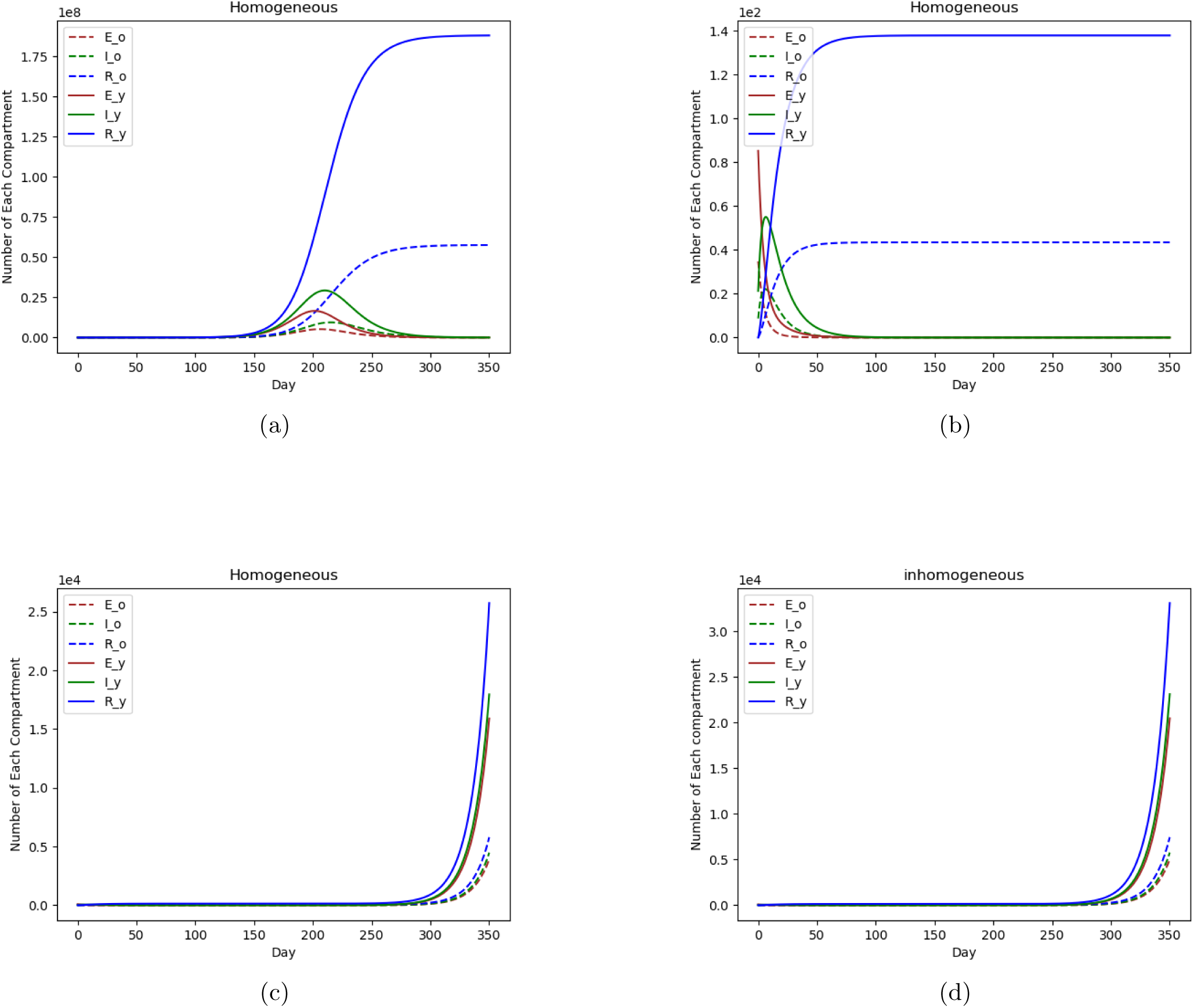
Epidemiological Change Over Time.

### Additional Sensitivity Analyses

#### Results of Sensitivity Analysis on Utility and Costs

The optimal policies of both the demographically homogeneous and inhomogeneous problems vary with different values of social benefits and costs in the objective function. We show some of these optimal policies in Table A.3.

In both homogeneous and inhomogeneous problems, the optimal policy remains unchanged when varying the costs of young and old patients in the range of 25-800 and 50-1600, respectively. The values of social benefits and costs in base case are given arbitrarily due to lack of related research on it, so it is compelling to verify the robustness (insensitivity) of optimal policy on benefits and costs.

**Table A.3:** Optimal policies under different benefits and costs.

| Social Benefit | Cost of Young Patient | Cost of Old Patient | Social Distancing Epochs in Time Homogeneous Policy | Social Distancing Epochs Time Inhomogeneous Policy |
| --- | --- | --- | --- | --- |
| 1 | 25 | 50 | 1, 2 | Y: 1, 2 O: N/A |
| 1 | 50 | 100 | 1, 2 | Y: 1, 2 O: N/A |
| 1 | 100 | 200 | 1, 2 | Y: 1, 2 O: N/A |
| 1 | 200 | 400 | 1, 2 | Y: 1, 2 O: N/A |
| 1 | 400 | 800 | 1, 2 | Y: 1, 2 O: N/A |
| 1 | 800 | 1600 | 1, 2 | Y: 1, 2 O: N/A |

Optimal policies under different costs and benefits. There are 5 epochs total (each epoch is 10 weeks long). The last two columns of the table give the epoch numbers in which the optimal policy recommends social distancing for either the whole population (homogeneous policy) or for the young or old cohorts (inhomogeneous policy)

Higher costs under the same optimal policy should result in lower daily utility per capita, but the difference is small (less than 0.01 utility units) as shown in Table A.4. The total sick weeks, total deaths, and the number of individuals recovered at end of time horizon do not change because the optimal policy and epidemiological parameters are the same even as the costs vary.

**Table A.4:** Comparison of outcomes under different costs; note that (100, 200) is the base case.

| Costs for (young, old): | (25, 50) |  | (50, 100) |  | (100, 200) |  | (200, 400) |  | (400, 800) |  | (800, 1600) |  |
| --- | --- | --- | --- | --- | --- | --- | --- | --- | --- | --- | --- | --- |
| Policy: | Homog. | Inhomog. | Homog. | Inhomog. | Homog. | Inhomog. | Homog. | Inhomog. | Homog. | Inhomog. | Homog. | Inhomog. |
| Average Daily Utility Per Capita | 0.60 | 0.72 | 0.60 | 0.72 | 0.60 | 0.72 | 0.60 | 0.72 | 0.60 | 0.71 | 0.60 | 0.71 |
| Young | 0.60 | 0.60 | 0.6 | 0.60 | 0.60 | 0.60 | 0.60 | 0.60 | 0.60 | 0.60 | 0.60 | 0.60 |
| Old | 0.60 | 1.00 | 0.60 | 1.00 | 0.60 | 1.00 | 0.60 | 1.00 | 0.60 | 1.00 | 0.60 | 1.00 |
| Total Sick Weeks Per 10,000 Persons | 1.50 | 1.97 | 1.50 | 1.97 | 1.50 | 1.97 | 1.50 | 1.97 | 1.50 | 1.97 | 1.50 | 1.97 |
| Young | 1.70 | 2.22 | 1.70 | 2.22 | 1.70 | 2.22 | 1.70 | 2.22 | 1.70 | 2.22 | 1.70 | 2.22 |
| Old | 1.10 | 1.36 | 1.10 | 1.36 | 1.10 | 1.36 | 1.10 | 1.36 | 1.10 | 1.36 | 1.10 | 1.36 |
| Total Deaths | 537 | 690 | 537 | 690 | 537 | 690 | 537 | 690 | 537 | 690 | 537 | 690 |
| Young | 41 | 53 | 41 | 53 | 41 | 53 | 41 | 53 | 41 | 53 | 41 | 53 |
| Old | 496 | 637 | 496 | 637 | 496 | 637 | 496 | 637 | 496 | 637 | 496 | 637 |
| Num. in Recovered Population at End of Time Horizon | 31,524 | 40,516 | 31,524 | 40,516 | 31,524 | 40,516 | 31,524 | 40,516 | 31,524 | 40,516 | 31,524 | 40,516 |
| Young | 25,730 | 33,067 | 25,730 | 33,067 | 25,730 | 33,067 | 25,730 | 33,067 | 25,730 | 33,067 | 25,730 | 33,067 |
| Old | 5,794 | 7,449 | 5,794 | 7,449 | 5,794 | 7,449 | 5,794 | 7,449 | 5,794 | 7,449 | 5,794 | 7,449 |

##### Clearance Rate Sensitivity Analysis

The optimal policies under different clearance rates are shown in table A.5. The optimal policy is sensitive to clearance rate in that higher clearance rates result in an optimal policy with less accumulated social distancing time.

**Table A.5:**
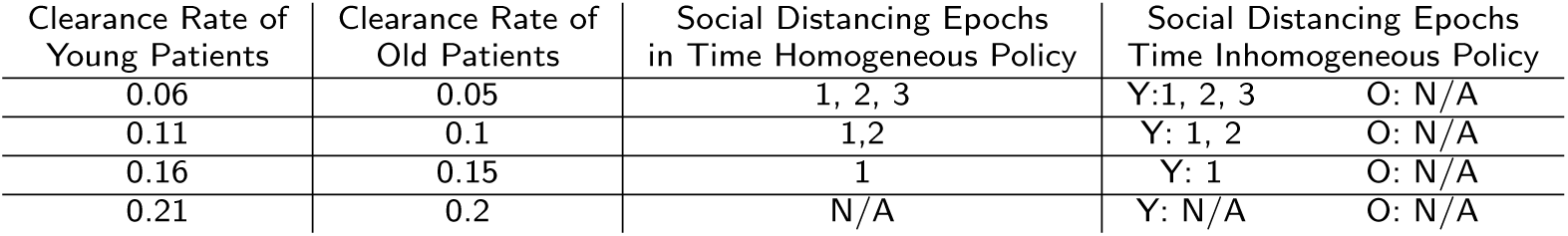
Optimal policies under different clearance rates

The improvement of clearance rates could increase social utility significantly (see Table A.6). When both clearance rates increase up to 0.2, the per-capita utility exceeds 0.9; note that a per-capita utility of 1 only happens when disease is eradicated and no social distancing occurs. In terms of health outcomes (see Table A.6), although the highest clearances rates could bring substantial social utility, it results in large sick-weeks per capita because the optimal policies recommend less social distancing.

**Table A.6:**
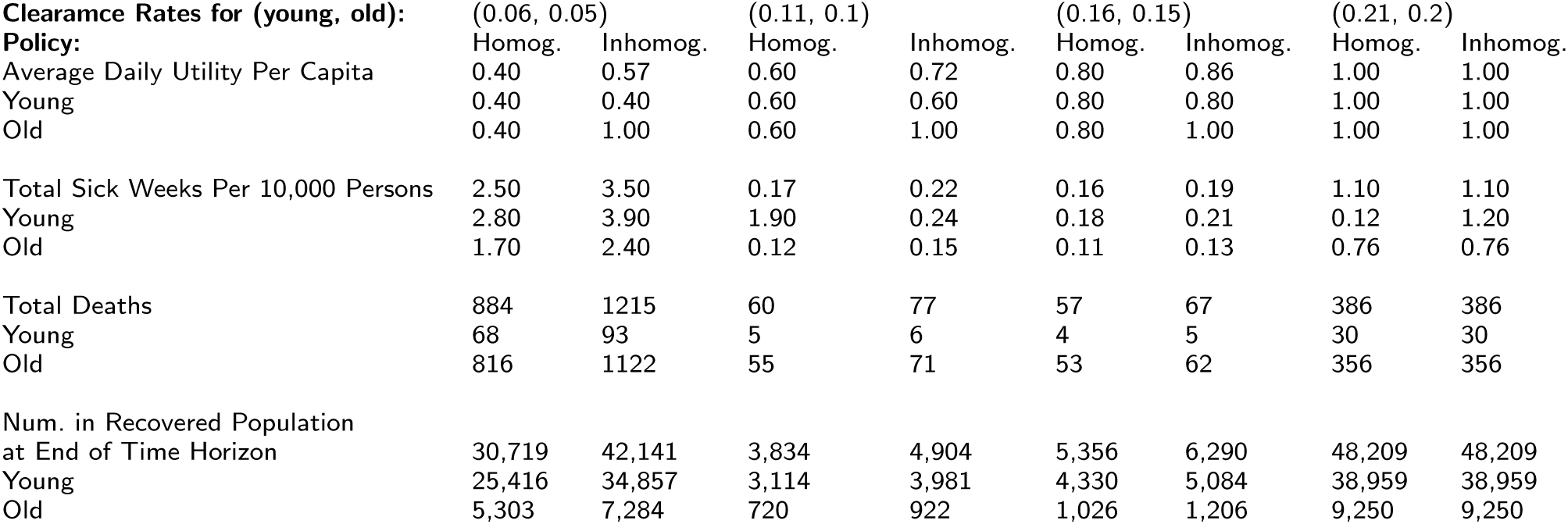
Comparison of outcome under different clearance rates.

#### Transmission Matrix Sensitivity

Betz et cl. studied 152 country’s population-based contacts by using a Bayesian hierarchical model to do estimation on the proclivity of age-and-location-specific contact patterns [24]. In their project, locations of contacts are divided into four locations, including home, school, work and other locations. The interval of each group by age is five years. We extract the American contact pattern data and summarize them in Table A.7. Each numeric entry in the table represents the contact rate within different age groups under each location. We generate transmission matricies using the procedure described in the main text. The complete estimated transmission rates are shown in Table A.9.

**Table A.7:**
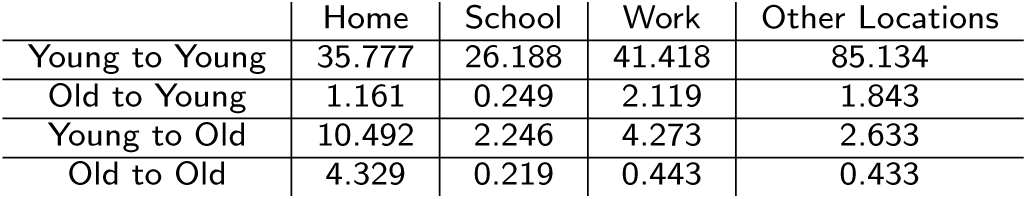
Average contact rates between different age groups under different locations.

|  | Home | School | Work | Other Locations |
| --- | --- | --- | --- | --- |
| Young to Young | 35.777 | 26.188 | 41.418 | 85.134 |
| Old to Young | 1.161 | 0.249 | 2.119 | 1.843 |
| Young to Old | 10.492 | 2.246 | 4.273 | 2.633 |
| Old to Old | 4.329 | 0.219 | 0.443 | 0.433 |

**Table A.8:**
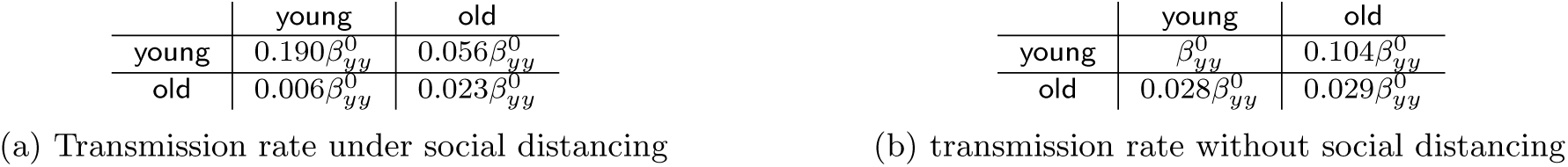
Ratios between base transmission rates and other transmission rates under new mixing contact matrix.

**Table A.9:**
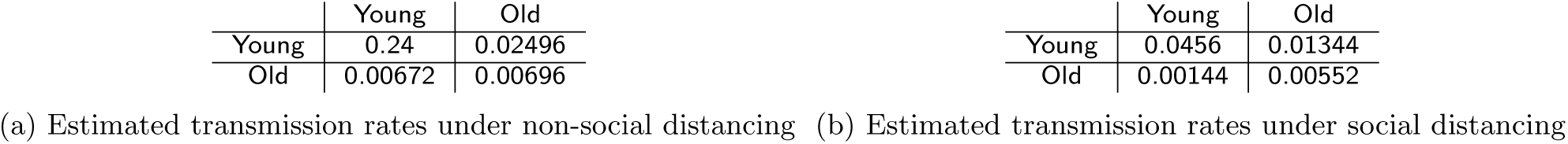
Estimated transmission rates under new mixing contact matrix.

The optimal inhomogeneous social distancing policy under these new estimated transmission rates is to socially distance at the second and third epoch for the young cohort while the old cohort never socially distance, which shifts social distancing one epoch later than the optimal policy derived from our base case contact pattern. The daily utility per capita derived from the new contact pattern is slightly lower than that of the one derived from the base case contact pattern (see Table A.10). The epidemiological change over time for homogeneous and inhomogeneous policies under the new pattern is shown in Figure A.5b and A.5c. Compared with Figures A.3c and A.3d, it would appear that we have similar epidemiological trends although more people would be infected in this new case. The deterioration in social utility and health outcomes under this new pattern is due to its larger estimated transmission rates under social distancing (see Tables 4 and A.9).

**Figure A.4:**
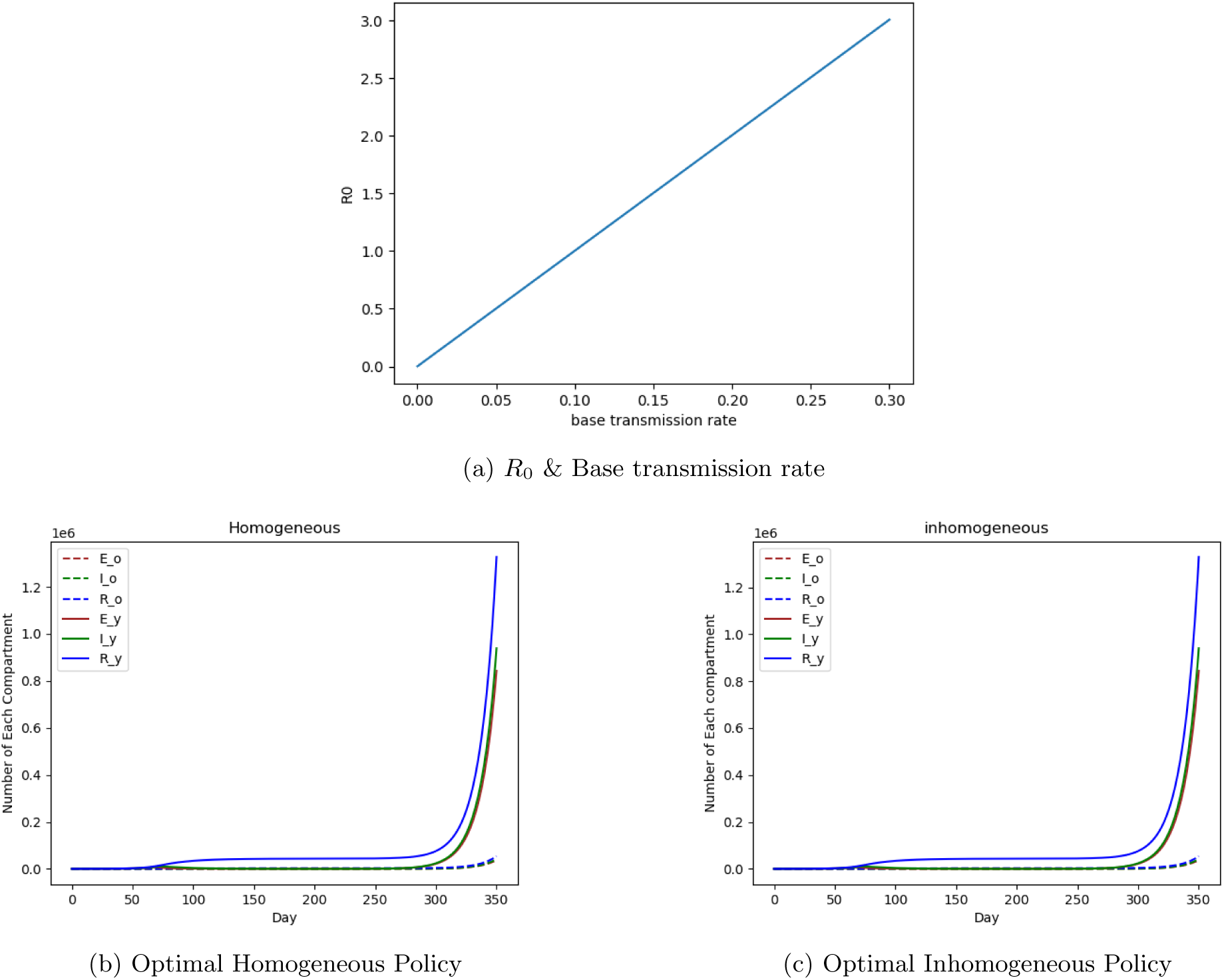
Epidemiological Change Over Time for the New Mixing Contact Matrix.

The higher average utility per capita earned by the inhomogeneous policy comes from the considerable increase of old’s average utility, who no longer social distance. Regarding health outcomes, the optimal policy under base case pattern (see Table A.10) outperforms the optimal policy under new mixing contact matrix because the transmission rates under social distancing estimated by new mixing contact matrix are larger (compare Tables 4 and A.9).

#### Mortality Rate Sensitivity Analysis

The results shown in Table A.11 reveal that the optimal policy is not sensitive to variation in mortality rates. Higher mortality rates result in lower number of sick-weeks but higher number of deaths, as expected.

**Table A.10:** Comparison of Outcomes Under Different Mixing Patterns.

| Costs for (young, old):<br>Policy: | Former Contact Pattern |  | New Contact Pattern |  |
| --- | --- | --- | --- | --- |
|  | Homog. | Inhomog. | Homog. | Inhomog. |
| Average Daily Utility | 0.60 | 0.72 | 0.59 | 0.70 |
| Young | 0.60 | 0.60 | 0.58 | 0.58 |
| Old | 0.60 | 1.00 | 0.60 | 1.00 |
| Total Sick Weeks Per 10,000 Persons | 1.50 | 1.97 | 6.60 | 66 |
| Young | 1.70 | 2.22 | 89 | 89 |
| Old | 1.10 | 1.36 | 10 | 10 |
| Total Deaths | 537 | 690 | 6765 | 6,781 |
| Young | 41 | 53 | 2123 | 2,127 |
| Old | 496 | 637 | 4642 | 4,654 |
| Num. in Recovered Population<br>at End of Time Horizon | 31,524 | 40,516 | 1.38M | 1.38M |
| Young | 25,730 | 33,067 | 1.33M | 1.33M |
| Old | 5,794 | 7,449 | 54,259 | 54,397 |

**Table A.11:** Optimal policies under different mortality rates.

| Mortality Rate of<br>Young Patient | Mortality Rate of<br>Old Patient | Social Distancing Epochs<br>in Time Homogeneous Policy | Social Distancing Epochs<br>Time Inhomogeneous Policy |  |
| --- | --- | --- | --- | --- |
| 0.00010 | 0.006 | 1, 2 | Y: 1, 2 | O: N/A |
| 0.00015 | 0.007 | 1, 2 | Y: 1, 2 | O: N/A |
| 0.00020 | 0.008 | 1, 2 | Y: 1, 2 | O: N/A |
| 0.00025 | 0.009 | 1, 2 | Y: 1, 2 | O: N/A |

**Table A.12:**
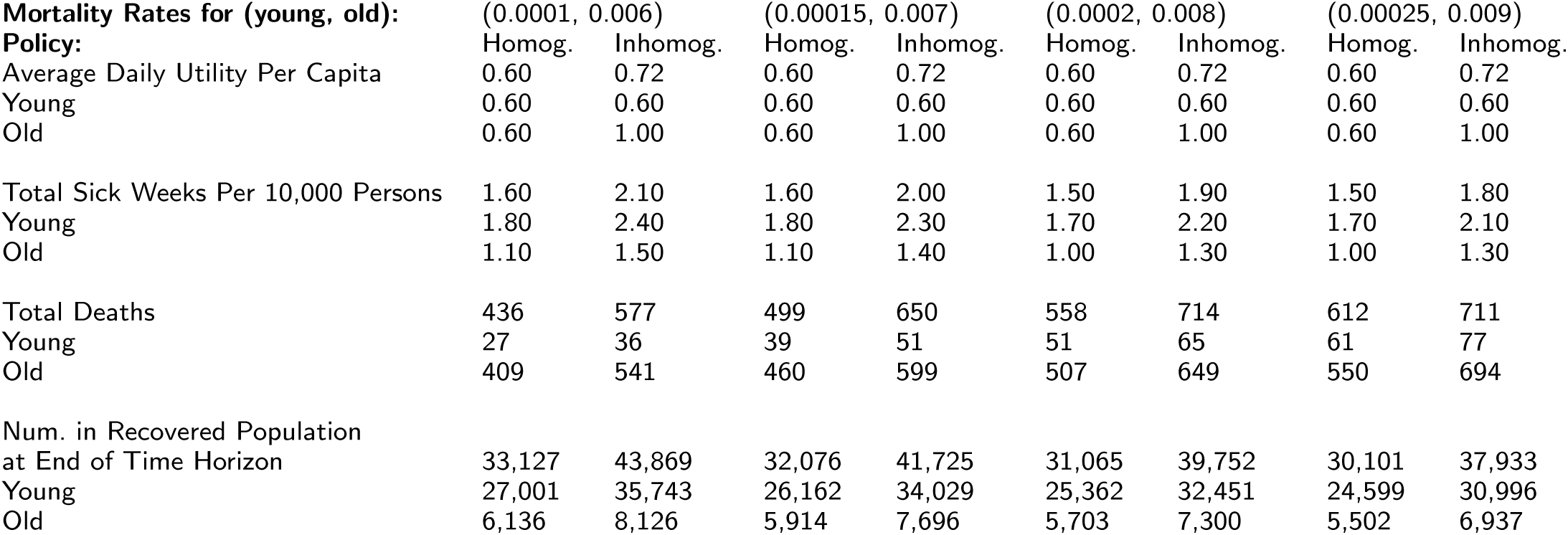
Comparison of outcome under different mortality rates.

| Mortality Rates for (young, old):<br>Policy: | (0.0001, 0.006) |  | (0.00015, 0.007) |  | (0.0002, 0.008) |  | (0.00025, 0.009) |  |
| --- | --- | --- | --- | --- | --- | --- | --- | --- |
|  | Homog. | Inhomog. | Homog. | Inhomog. | Homog. | Inhomog. | Homog. | Inhomog. |
| Average Daily Utility Per Capita | 0.60 | 0.72 | 0.60 | 0.72 | 0.60 | 0.72 | 0.60 | 0.72 |
| Young | 0.60 | 0.60 | 0.60 | 0.60 | 0.60 | 0.60 | 0.60 | 0.60 |
| Old | 0.60 | 1.00 | 0.60 | 1.00 | 0.60 | 1.00 | 0.60 | 1.00 |
| Total Sick Weeks Per 10,000 Persons | 1.60 | 2.10 | 1.60 | 2.00 | 1.50 | 1.90 | 1.50 | 1.80 |
| Young | 1.80 | 2.40 | 1.80 | 2.30 | 1.70 | 2.20 | 1.70 | 2.10 |
| Old | 1.10 | 1.50 | 1.10 | 1.40 | 1.00 | 1.30 | 1.00 | 1.30 |
| Total Deaths | 436 | 577 | 499 | 650 | 558 | 714 | 612 | 711 |
| Young | 27 | 36 | 39 | 51 | 51 | 65 | 61 | 77 |
| Old | 409 | 541 | 460 | 599 | 507 | 649 | 550 | 694 |
| Num. in Recovered Population<br>at End of Time Horizon | 33,127 | 43,869 | 32,076 | 41,725 | 31,065 | 39,752 | 30,101 | 37,933 |
| Young | 27,001 | 35,743 | 26,162 | 34,029 | 25,362 | 32,451 | 24,599 | 30,996 |
| Old | 6,136 | 8,126 | 5,914 | 7,696 | 5,703 | 7,300 | 5,502 | 6,937 |

